# Is Rapid Molecular Testing enough In Screening Resistant Tuberculosis In Community

**DOI:** 10.1101/2021.02.25.21252270

**Authors:** Dhaker Ashok, Bahal Ashish, Mangal Vishal, K Yadav Arun, Singhal Anuj, Gupta Anoushka

## Abstract

**Background:** The study aimed to compare the sensitivity and specificity of cartridge based nucleic acid amplification test (CBNAAT) for diagnosis of Drug-Resistant tuberculosis (DRTB) with culture sensitivity assays.

**Methods:** Patients with cough symptoms for more than two weeks with any one symptom such as night sweats, fever, and unintentional weight loss were enrolled. Cases where *Mycobacterium Tuberculosis* was detected on sputum CBNAAT, were included in the study. Demographic variables, clinical features, and chest radiographs were collected. Each sputum sample was divided into three aliquots: smear microscopy, culture, and genotypic drug sensitivity testing (DST). Results of all three diagnostic modalities were compared with CBNAAT.

**Results:** Out of 236 patients with sputum positive CBNAAT, 49.4 % (117/236) were rifampicin-resistant while 50. 6 % (119/236) were Rifampicin sensitive. The genotypic DST assays carried out on all enrolled patients showed that 76. 3 % (181/236) patients were resistant to one or more first-line or second-line antitubercular (ATT) drugs, while 23.7 % (55/236) patients were sensitive to all ATT drugs. On concordant analysis of CB NAAT with DST assays, we found that among 119 CB NAAT rifampicin sensitive patients, 66 patients were resistant to first-line or second-line antitubercular drugs.

**Conclusion:** This study found that the screening of DRTB with CBNAAT at the community level is suboptimal compared to the gold standard. Although CBNAAT’s sensitivity in detecting DRTB is significantly higher, the specificity is lower in that population who have received ATT earlier.

## Introduction

Drug-Resistant Tuberculosis (DR TB) remains a significant public health concern in many countries. There is a trend for increasing Multi Drug-Resistant Tuberculosis (MDR TB) cases as a proportion of all globally notified tuberculosis cases. The incidence rate of MDR TB in India is 9.6 %. ^[1]^

Cartridge Based Nucleic Acid Amplification Test (CB-NAAT) is an automated cartridge-based molecular technique based on Polymerase Chain Reaction (PCR), which can detect Mycobacterium Tuberculosis (MTB) from any of the pulmonary or extrapulmonary specimens with varying sensitivity & specificity.^[2]^ The test carries the advantage that it gives rapid results and can detect rifampicin resistance (RRDR of rpo B5 allele) within two hours.^[3]^ Gene Xpert is the only rapid test for TB diagnosis currently recommended by WHO to detect M*ycobacterium tuberculosis* and report rifampicin resistance directly from sputum samples.^[4-6]^The reported sensitivity ranges between 72.5–98.2 % (smear-negative and smear-positive samples, respectively) with a specificity of 95 %.^[4]^ However, Drug Susceptibility Test (DST) remains the gold standard for the detection of drug resistance as it can detect drug-resistance against any of the First Line Anti Tubercular Therapy (FL ATT) or Second Line Anti Tubercular Therapy (SL ATT).^[5]^

Rifampicin resistance (RR) was considered as a predictor of multidrug-resistant TB (MDR-TB) because 78 % of RR-TB were MDR -TB. ^[6]^ Genotypic studies have shown that *Kat G P ser 315*, mediated isoniazid resistance, is among the earliest to evolve even before rifampicin resistance and is related to mycobacterium’s virulence.^[7]^ CBNAAT can quickly detect common mutations in the 81 base pair rifampicin Resistance Determining Region (RRDR). The rpoB mutations outside the RRDR are undetectable by CBNAAT, the most common being *Ile 491 Phe mutation* in the *rpo B* gene present in 56% of rifampicin resistant TB cases.^[8]^It can detect rifampicin resistance only if *the rpo B allele* responsible for Rif resistance is present in at least 65% of DNA present in the sample.^[9]^ Similarly, CB NAAT has a low sensitivity to detect *Leu 533 CCG, 572 Phe& 531 CTG mutation*s responsible for up to 5% of all Rif resistance.^[9-11]^

We studied the performance, validity & accuracy of CB-NAAT for accessing drug resistance among pulmonary TB compared with the solid culture DST.

## Material and Methods

The study was conducted at the Directly Observed Treatment Centre (DOTS) nodal center affiliated with a tertiary care Governmental hospital of western Maharashtra. All presumptive adult pulmonary TB patients reporting at DOTS Centre from Oct 2018 to Sep 2020 were enrolled in the study period.

Participants of either sex aged between 15 to 70 years, positive for sputum CB NAAT and not on ATT past two months, were enrolled for the study after taking informed written consent.

Patients with cough symptoms for more than two weeks with any one symptom such as night sweats, fever, and unintentional weight loss were enrolled. The sputum samples were collected for rapid molecular testing. Cases where MTB was detected on sputum CBNAAT (Rifampicin sensitive or Rifampicin resistant) were included. Previously diagnosed drug-resistant Pulmonary TB by sputum CBNAAT having constitutional symptoms but not on any ATT for a minimum of two months duration, who reported at DOTS center for routine follow up were also included in the study. Patient information, including age, immune surveillance status, clinical features, and chest radiographs, were collected. Each sputum sample was divided into three aliquots: smear microscopy, Liquid culture (LC) by BACTEC (Becton Dickinson Microbiology Systems, Cockeysville, Md.), and genotypic DST. Results of all three diagnostic modalities were compared with CBNAAT reports. The study was approved by Institutional Ethics Committee vide letter no: 1 IEC / oct / 2018 dated 22 Oct 18.

The sample size was calculated apriori to estimate the 95 % confidence interval for sensitivity & specificity of CB NAAT in comparison with DST with 5% absolute error of margin. Following statistical formula was used to establish sample size.

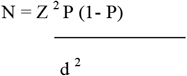

where P = Expected prevalence or proportion, d = precision and Z = statistic for level of confidence (in our case 1.96 as per 95 % CI), using the test sensitivity as 92% and specificity 95% (12). The calculated sample size was 186.

Data was collated in an excel sheet. Continuous variables were described as mean and standard deviation, and categorical variables were defined as numbers and percentages. Sensitivity and specificity were calculated. A p-value of less than 0.05 was considered statistically significant. All data were analyzed using SPSS statistical software version 25.0(IBM, USA).

## Results

Our study screened 496 patients who had constitutional symptoms of low-grade fever, chronic cough, and weight loss of more than two weeks or previously diagnosed Pulmonary TB by CBNAAT but with a drug default of more than two months.

Two hundred thirty-six patients were included in the study with a positive CBNAAT report (Rifampicin sensitive or Rifampicin resistant). Among the majority of the patients were middle-aged, with 33.1% patients (n= 78) in their fourth decade and 24.6 % patients (n=58) in their fifth decade. Out of total enrolled patients 61.4% (n=145) were males. 177/236 patients were newly diagnosed pulmonary tuberculosis, whereas 59/236 patients were default cases.

Out of 236 patients, thirty patients were HIV positive, while 141 were HIV negative. Data about the rest of the patients was unknown. Further evaluation of HIV-positive patients revealed that 80 % (n=24) were MDR TB cases detected either by DST or CBNAAT.

Chest radiographs of all the patients enrolled were studied. Variations in the pattern of chest radiograph among drug-sensitive and DR TB cases are depicted in **Fig 1**. Out of 180 DR TB by DST assays, 54.4 % (n=98) patients had cavitary lesions on CXR. However out of 56 drug-sensitive cases five percent (n=3) had lung collapse, 17 % (n=10) had consolidation, 12 % (n=7) had pleural effusion and five percent (n=3) had cavitation.

**Figure 1.**
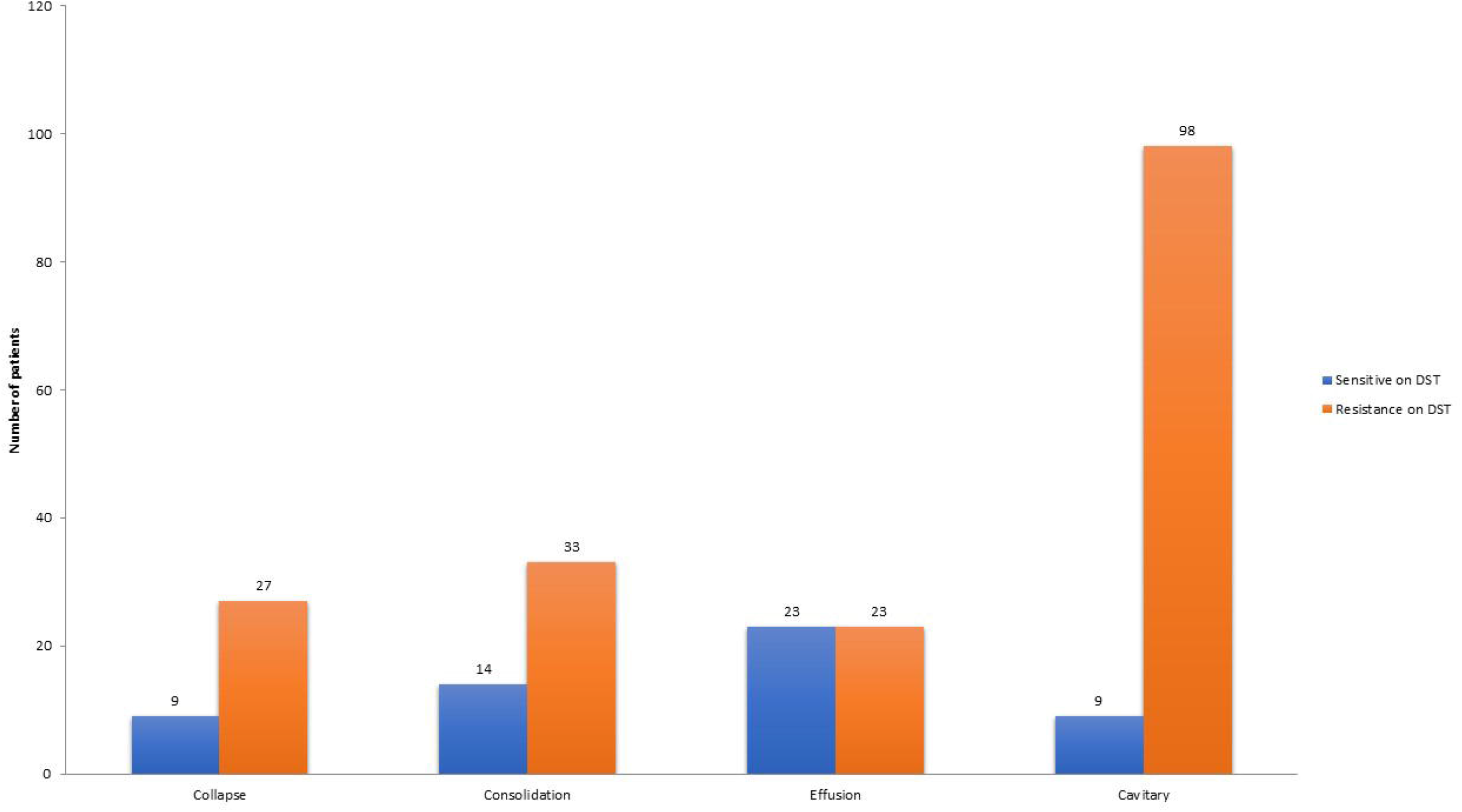
Chest radiographs patterns among drug resistant and sensitive pulmonary tuberculosis patients

Concordant analysis between DST and sputum smear microscopy revealed that microscopy positivity was more among drug-resistant cases (67 %; 121/180 samples) than drug-sensitive cases (21.4 %; 12/56 samples). In our study overall sensitivity of sputum microscopy to detect *Mycobacterium Tuberculosis* was found to be 56.4 % (133 among 236).

Out of these 236, patients with sputum positive CBNAAT (n= 236), 49.4 % (n= 117) were Rifampicin Resistant (RR) while 50. 4 % (n= 119) were rifampicin sensitive. The genotypic DST assays carried out on all enrolled patients showed that 76.3 % (n= 181) patients were resistant to one or more FL ATTs or SL ATTs while 23.7 % (n= 55)patients were sensitive to all ATTs. Among all the study participants 56.4 % (n=133) patients had sputum smear positive by ZN stain while 88.6 % (n=209)showed growth on Liquid culture (BACTEC) media. The details are placed in (**Table 1a and 1b**)

**Table 1a.** Distribution of all test results.

| **Tests** | **Samples taken for comparative analysis ( n=236 ), n ( % )** |  |
| --- | --- | --- |
|  | **Positive** | **Negative** |
| **Microscopy** | 133 ( 56.4 ) | 103 ( 43.6 ) |
| **Liquid Culture** | 209 ( 88.6 ) | 27 ( 11.4 ) |
| **CBNAAT** | 236 ( 100 ) |  |
| **DST** | 236 ( 100 ) |  |

CBNAAT- Cartridge based nucleic acid amplification test, DST- Drug sensitivity testing

**Table 1b.** Distribution of test results for Drug-resistant tuberculosis.

| **Tests** | **Samples taken for comparative analysis ( n=236 ),** |  |
| --- | --- | --- |
|  | **n (%)** |  |
|  | **Sensitive** | **Resistant** |
| **CBNAAT** | 119 (50 .4) | 117 (49.6) |
| **DST** | 56 (23.7) | 180 (76.3) |

CBNAAT- Cartridge based nucleic acid amplification test, DST- Drug sensitivity testing

An absolute number of DR TB identified by CBNAAT and DST with simultaneous analysis of sputum smear and Liquid Culture (LC) reports is depicted in **Table 2a** and **Table 2b**, respectively. Most of the discordance among results of CBNAAT and DST was found in the sub-group of patients showing positive reports for both sputum smear microscopy and LC.

**Table 2a.** Performance of CBNAAT among various target groups.

| **Microscopy** | **Liquid Culture** | **CBNAAT** |  | **Total** | **P-value** |
| --- | --- | --- | --- | --- | --- |
|  |  | **Sensitive n (%)** | **Resistant n (%)** |  |  |
| Negative | Negative | 24 (88.9) | 03 (11.1) | 27 | 0.014 |
|  | Positive | 47 (61.8) | 29 (38.2) | 76 |  |
| Positive | Positive | 48 (36.1) | 85 (63.9) | 133 |  |
| Total |  | 119 | 117 | 236 |  |

CBNAAT- Cartridge based nucleic acid amplification test, Significant P value <0.05

**Table 2 b:** Performance of DST among various target groups.

| Microscopy | Liquid Culture | DST |  | Total | P-value |
| --- | --- | --- | --- | --- | --- |
|  |  | Sensitive n (%) | Resistant n (%) |  |  |
| Negative | Negative | 21 (77.8) | 06 (22.2) | 27 | <0.001 |
|  | Positive | 23 (30.3) | 53 (69.7) | 76 |  |
| Positive | Positive | 11(8.3) | 122 (91.7) | 133 |  |
| Total |  | 55 (23.3) | 181 (76.7) | 236 |  |

DST- Drug Sensitivity testing, Significant P-value <0.05

A concordant analysis of CBNAAT results was done with DST assays. Among 119 CB NAAT rifampicin-sensitive patients, 66 patients were DR TB to any of the FL or SL ATTs. Thirty six percent (n=24) were relapse cases while rest 64 % (n=42) were newly detected. Further analysis showed that out of these 66 DR TB patients, 71.2 % (n=47) were resistant to one or more SL ATTs (maximum resistance to Moxifloxacin 40.9 %) while only 28.8 % (n=19) were resistant to only FL ATTs. Isolated Isoniazid resistance with Rifampicin susceptible (Hr TB) was found among 15 patients, and isolated Rifampicin Resistance (RR TB) for FL ATTs was detected among 27 patients. (**Table 3a**)

**Table 3a.** Pattern of Antitubercular drug resistance among Rifampicin sensitive CBNAAT samples

| **Anti-Tubercular Drug** | **Samples taken for comparative analysis (n= 236), n (%)** |  |  |
| --- | --- | --- | --- |
|  | **Sensitive** | **Resistance** | **Total** |
| **Streptomycin** | 107 (89.9) | 12 (10.1) | 119 |
| **Isoniazid** | 97 (81.5) | 22 (18.5) | 119 |
| **Rifampicin** | 84 (70.6) | 35 (29.4) | 119 |
| **Ethambutol** | 113 (95.0) | 6 (5.0) | 119 |
| **Kanamycin** | 96 (80.7) | 23 (19.3) | 119 |
| **Capreomycin** | 108 (90.8) | 11 (9.2) | 119 |
| **Moxifloxacin** | 92 (77.3) | 27 (22.7) | 119 |

**Table 3b.**
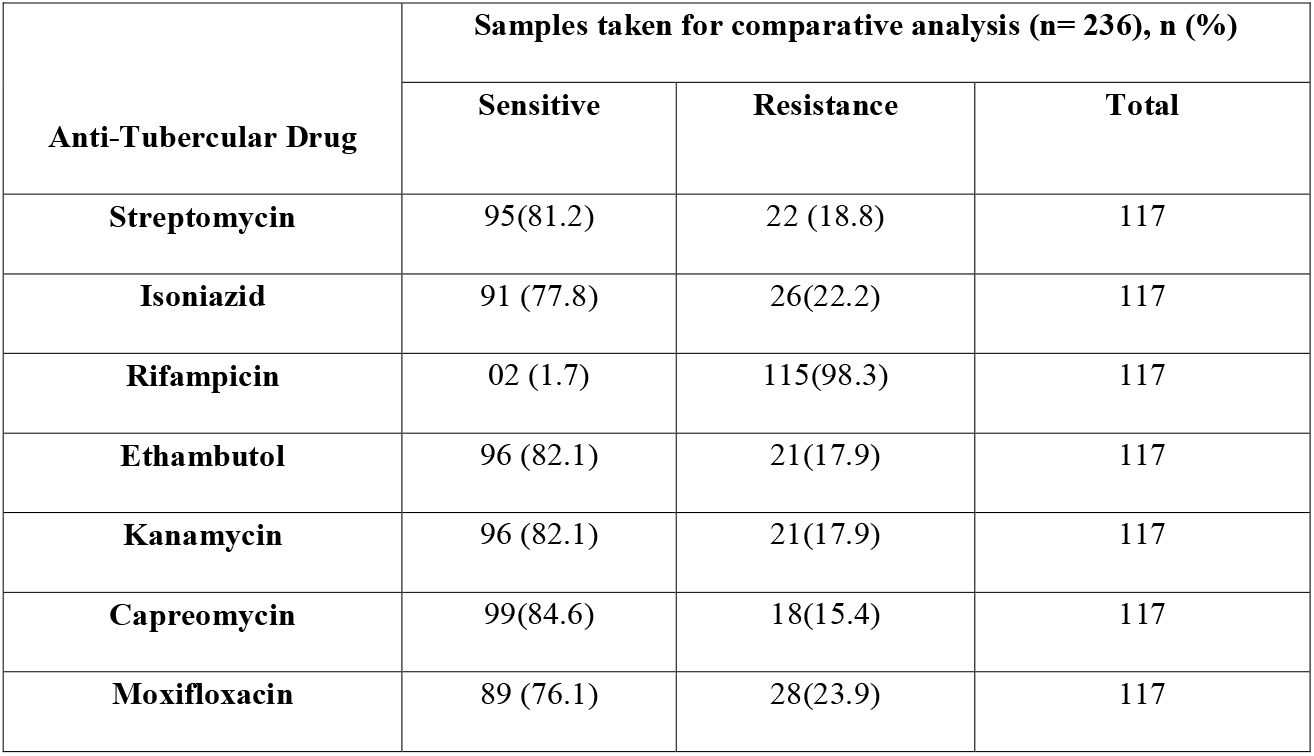
Pattern of Antitubercular drug resistance among Rifampicin resistant CBNAAT samples

In the second group among 117 CB NAAT rifampicin-resistant patients, 115 were DR TB on DST to various FL and SL ATTs, of which 27.8 % (n= 32) were relapse cases while the rest 72 .2 % (n= 83) were newly detected. Patients resistant to one or more SL ATTs in this group were only 33.3 % (n= 39). **(Table 3 b)** None of the cases was found to be Hr TB or RR TB in this group.

The sensitivity, specificity, PPV & NPV of CB NAAT for detecting rifampicin resistance on sputum for pulmonary TB compared with the gold standard DST assays was 97.67 %, 76.67 %, 70.59 %, and 98.29 %, respectively.

Drug resistance patterns for various FL and SL ATTs were compared among newly detected pulmonary TB and relapse cases. In our study,75 % (n=177) of patients were newly detected pulmonary TB who were recruited before initiation of treatment, while the remaining25 % (n=59) patients were cases of pulmonary TB with more than two months of ATT default or relapse TB. In the first group of 177newly detected pulmonary TB cases, DST assays revealed that70.6 % (n= 125) patients were DR TB while the remaining 29.4 % (n= 52) were sensitive to all ATTs. Among newly detected pulmonary TB cases found to be DR TB,65.6 % (n= 82) patients were isolated RR TB, and 28 % (n= 35) were resistant to one or more SL ATTs. Most of the resistance to SL ATTs was attributed to moxifloxacin (n= 25). In the other group comprising 59 relapse pulmonary TB cases, 94.9 % (n= 56) were DR TB, which was statistically significant (p-value < 0.001). Among this group, only one patient was isolated RR TB, and 91 % (n= 51) were resistant to SL ATTs. The comparison of resistance to different anti-tubercular drugs amongst the rifampicin sensitive and rifampicin resistant samples on CBNAAT is depicted in **Figure 2**.

**Figure 2.**
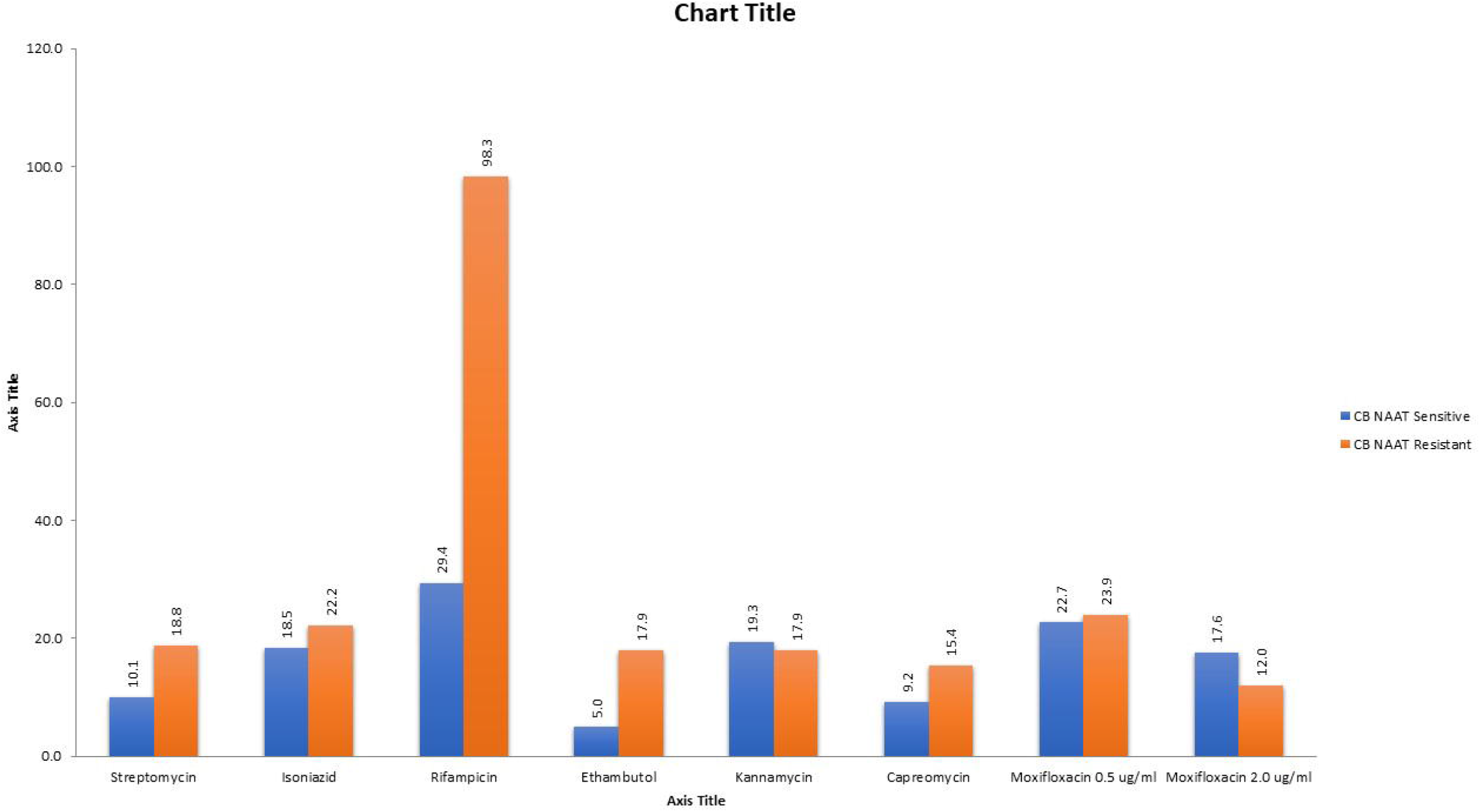
The comparison of resistance to different anti-tubercular drugs amongst the rifampicin sensitive and rifampicin resistant samples on CBNAAT

## Discussion

The main gap in our fight against TB is the lack of a fast, accurate & economic test that can be used in a resource-limited setting. The development of nucleic acid-based tests like CBNAAT has provided novel avenues for generating highly sensitive point-of-care tests but still has significant limitations in diagnostic accuracy for DR TB.^[12]^

Our study found that DR TB was more common among relapse cases. These findings corroborated with earlier studies which also substantiated that drug-resistant TB was more common among relapse cases than freshly detected TB cases.^[13-14]^ This observation was likely due to the development of antibiotic resistance caused by the low serum concentration of antitubercular drugs and partly due to cross-resistance among antitubercular drugs. Poor drug compliance among patients on long-term treatment also comes as contributing factor. The study also found a difference in drug resistance patterns in newly detected and relapse cases. While the newly detected drug-resistant pulmonary TB cases were mostly isolated rifampicin-resistant TB cases, in contrast, most of the relapsed drug-resistant TB cases were resistant to second-line ATTs.

Our study found the specificity of CBNAAT to detect drug-resistant cases of pulmonary TB was 76 %, which was less compared to previous studies. ^[4,15]^A significant proportion of drug-resistant TB missed by CBNAAT (Rif sensitive) were relapsed cases (36% vs. 27. 8 %). Twenty-five percent of our study population included relapse cases that were already exposed to ATT earlier. Our study recommends that a high suspicion of MDR TB should be placed in relapse pulmonary TB cases. There is a perceived high probability of them harboring drug-resistant TB apart from isolated rifampicin resistance. This could have been the reason for the low specificity of CBNAAT found in our study. Our study also found that the resistance to second-line ATTs was more common among Rifampicin sensitive CBNAAT than Rifampicin Resistance CBNAAT (71.2 % vs. 33.3 %). This was likely because Rifampicin sensitive CBNAAT reports were either false negative or patients were resistant to other second-line ATT without concurrent Rif resistance like HR TB.

In our study, 80 % of HIV seropositive were found to be MDR TB cases. This corroborated with the findings of a previous study done by Sunil Sethi et al^[16]^, which stated a significantly higher association of MDR TB with HIV seropositive patients (27.3%) as compared to HIV seronegative. The proportion of similar cases was higher in our study, which could have been due to referral bias as our study was conducted at a tertiary care referral center. Our study’s findings were also analogous to a study conducted by Luz et al., which showed that a high proportion of cavitary lesions were observed among MDR TB cases.^[14]^ Cavitary lesions on Chest X-ray may be used as a soft indicator of MDR TB.

Previous studies have demonstrated that the global average of Isoniazid resistant TB was 11.6 % in new TB cases & 7.2 % in previously treated TB cases.^[17]^ This study found 15 patients with isolated Isoniazid resistance without Rifampicin resistance (Hr TB) on DST. Among these 4.6 % (n=11) were new TB cases and 1.6 % (n=4) were previously treated TB cases. The findings were in concurrence with the previous study.

Similarly, the resistance pattern to other SL ATTs was also studied. Thirty percent (n= 55) of DR TB cases were found to have resistance to Fluoroquinolones (FQ) with or without evidence of concurrent resistance to First-line ATT on DST. Previous studies have also verified that the prevalence of resistance to Levofloxacin was comparatively more in South Asian countries.^[14]^This could be due to the non-judicious use of FQ in the subcontinent.

This study’s limitations were mainly due to the lack of sufficient clinical data available on subjects and their follow up during treatment, which would have helped us identify other predictors of DR TB’s emergence. Future work should include clinical data, including atypical presentations in high-risk groups like children & the elderly, together with longitudinal studies. CBNAAT cannot quantify Rif Resistance, which is a valuable tool in TB endemic settings. A significant number of Rifampicin sensitive cases as per CBNAAT were found to have either isolated Isoniazid mono resistance or resistance to second-line ATT on DST assays. CBNAAT cannot be used for assessing the emergence of Rifampicin resistance during treatment.^[4]^ It cannot detect Hetero resistance. It gives false-positive results in mixed TB infections.

We need some alternate tests to overcome these fallacies of CBNAAT. These can be corrected by the more aggressive introduction of MDR-TB diagnostic tests like Line Probe Assay (LPA) at primary health care setups and utilizing Whole Genome Sequencing for mutational analysis of drug-resistant strains. The frequency of 533Pro mutation responsible for low-Level Resistance (LLR) has been reported globally to range from 3% to 6% and hence may not be as salient as the other more frequent high-level resistance mutations.

## Conclusions

This study found that the rapid molecular technique in screening DRTB at the community level is suboptimal compared to the gold standard solid cultures method. This study highlights the high prevalence of MDR-TB cases in patients visiting Governmental TB institutes with relapse TB. Although the sensitivity of CBNAAT in the detection of DR pulmonary TB is significantly higher, the specificity is lower in a population that comprises a high number of cases of those who have received ATT earlier. Our study recommends that Immuno-surveillance patients not responding to usual first-line ATT should have a high suspicion of acquiring MDR TB strain and accordingly investigated.

### Recommendations of this Study

The inclusion of specific mutant probes in the CBNAAT design could assist treatment decisions. A suitable alternative test will minimize the number of false-negative drug-resistant TB cases being treated inappropriately for several weeks before obtaining the DST results. This will also cease the potential spread of RR /MDR-TB pathogen to many public places, causing grave economic & public burden.

## Data Availability

All the data is available with the authors on request

## Acknowledgement

We acknowledge the help rendered by Dr Sanjay Darade, District Tuberculosis Officer, Pune for unstinted support during this study. This study was conducted as part of MUHS MD degree award in subject of General Medicine. The authors thank all study participants, TB diagnostic laboratory staff and TB Clinic staff of Nodal Centre at our institute.

